# High prevalence and risk of malaria among asymptomatic individuals from villages with high rates of artemisinin partial resistance in Kyerwa district, North-western Tanzania

**DOI:** 10.1101/2023.10.05.23296564

**Authors:** Salehe S. Mandai, Filbert Francis, Daniel P. Challe, Misago D. Seth, Rashid A. Madebe, Daniel A. Petro, Rule Budodo, Angelina J. Kisambale, Gervas A. Chacha, Ramadhan Moshi, Ruth B. Mbwambo, Dativa Pereus, Catherine Bakari, Sijenunu Aaron, Daniel Mbwambo, Abdallah Lusasi, Stella Kajange, Samuel Lazaro, Ntuli Kapologwe, Celine I. Mandara, Deus S. Ishengoma

## Abstract

**Background:** Tanzania adopted and has been implementing the World Health Organization (WHO) recommended interventions to control and eventually eliminate malaria. However, malaria is still a leading public health problem and the country experiences heterogeneous transmission; but the drivers of these patterns are not clearly known. This study assessed the prevalence and risk of malaria infections among asymptomatic individuals living in a hyperendemic area which has high prevalence of artemisinin partial resistant parasites in Kyerwa District of Kagera region, North-western Tanzania.

**Methods:** This was a community-based cross-sectional survey that recruited participants from five villages of Kyerwa district in Kagera region. Demographic, anthropometric, clinical, parasitological, types of houses inhabited and socio-economic status (SES) data were collected using electronic capture tools running on Open Data Kit (ODK). Risk factors associated with malaria infections were determined by univariate and multivariate logistic regression and the results were presented as crude (cOR) and adjusted Odds Ratio (aOR), with 95% confidence intervals (CI).

**Results:** A total of 4,454 individuals were tested using malaria rapid diagnostic tests (RDTs) and 1,979 (44.4%) had a positive test. The prevalence of malaria varied from 14.4% to 68.5% with significant differences among the villages (p<0.001). The prevalence and risk of malaria infections were significantly higher in males (aOR =1.25, 95% CI: 1.06 - 1.48, p=0.04), school children ((aged 5 – 10 years, aOR =4.09, 95% CI: 3.39 – 5.10, p<0.001) and (10-15 years, aOR=4.40, 95% CI: 3.46 – 5.59, p<0.001)) and among individuals who were not using bed nets (aOR =1.29, 95% CI: 1.10 – 1.42, p=0.002). Other risks of malaria infections included lower SES (aOR=1.27, 95% CI:1.04 – 1.55, p<0.001) and living in houses with open windows (aOR=1.26, 95% CI: 1.03 −1.54, p=0.024).

**Conclusion:** This study showed high prevalence of malaria infections and high heterogeneity at micro-geographic levels. The risk of malaria infections was higher in school children, males, individuals who did not use bed nets, and among participants with low SES or living in poorly constructed houses. These findings provide important baseline data in an area with a high prevalence of artemisinin partial-resistant parasites and will be utilized in future studies to monitor the trends and potential spread of such parasites.

## Background

In the past two decades, there have been enhanced malaria control efforts which resulted in a significant decline of the disease burden globally, but progress has stalled since 2015 [1]. According to the World Health Organization (WHO), there were an estimated 247 million malaria cases and 619,000 deaths in 2021 [2]. The WHO African Region (WHO-Afro), is the leading region with the highest malaria cases and deaths and accounted for 95% of the cases and 96% of malaria deaths in 2021, with an estimated 234 million cases and 593,000 deaths [2]. Although the global scale-up of effective malaria interventions has saved millions of lives globally and cut malaria mortality by 36% from 2010 to 2020, the resulting optimism leading to hopes and plans to eliminate and ultimately eradicate malaria are facing imminent threats. In Tanzania, over 93% of the population lives in areas where transmission occurs [3]. The country has recently transitioned from very high and stable to areas with varying transmission intensities [4,5]. Like in many malaria-endemic countries in the WHO-Afro region, *Plasmodium falciparum* is the cause of most of the infections in Tanzania, with other species being *Plasmodium ovale* spp and *Plasmodium malariae* [6–8]. Although *Plasmodium vivax* has been reported in a few studies in Tanzania [6,8], its public health significance remains unknown. The biggest contemporary challenges to malaria elimination include insecticide resistance by malaria vectors [9,10], emergence and spread of drug resistance [11,12], emergence of parasites that cannot be detected by the rapid diagnostic tests (RDTs) due to deletions of the histidine-rich protein 2/3 (hrp2/3) gene [13,14] and emergence as well as spread of invasive *Anopheles stephensi* vectors [15,16]. There are also non-biological threats such as reduced funding for malaria control and political will at global and local/country levels [17,18].

To accelerate progress towards its elimination targets by 2030, Tanzania adopted WHO’s recommendations and has been implementing effective interventions over the past two decades to control and eventually eliminate malaria. The Tanzanian National Malaria Control Program (NMCP) has endeavoured to ensure high coverage and use of such interventions, focusing on vector control, case management and preventive therapy. Vector control interventions include the use of insecticide treated nets (ITNs) and/or long-lasting insecticidal nets (LLINs), indoor residual spraying (IRS) and larval source management (LSM) while for effective case management, NMCP uses RDTs and artemisinin-based combination therapy (ACT). To prevent malaria in pregnancy, NMCP deployed and strives to increase the proportion of women receiving two or more doses of sulphadoxine-pyrimethamine (SP) for intermittent preventive treatment (IPTp) of malaria during pregnancy [19]. These intervention yielded positive results and a declining trend of malaria burden was reported as shown by the decline of prevalence from 18% in 2008 to 7.3% in 2017 [20].

Based on recent reports, malaria remains a major public health problem and the country still experiences persistent transmission in some regions [2], but the drivers of these patterns are not clearly known. Malaria transmission in Tanzania has become heterogenous with high burden in north-western, southern and western regions while the central corridor, north-eastern and south-western parts of the country have low to very low transmission intensities [21,22]. The recent epidemiological transition and trends of malaria have raised critical questions which need to be addressed to allow the country to progress to its elimination targets by 2030. In addition, Tanzania faces emerging and endemic challenges such as low coverage of existing interventions, emergency and spread of drug and insecticide resistance and weakness of the public health system [23]. Most of these challenges are particularly context specific and require innovative and multi-faced approaches to ensure high impacts are attained and sustained. The interventions should also target and aim to eliminate the remaining pockets in low transmission areas as well as reducing the disease burden in high transmission areas and dealing with populations at higher risk [24]. Thus, NMCP needs to strategically and innovatively continue to implement the WHO recommended interventions including upgrading and making malaria surveillance a core intervention to facilitate the ongoing elimination efforts in all areas based on the local burden and level of transmission [25].

Tanzania faces biological and other threats which may compromise its progress and prospects to eliminate malaria by 2030 [26]. Of the biological threats, Tanzania has reported high prevalence of mosquitoes with resistance to multiple insecticides [10] and it is at high risk of the invasive *Anopheles stephensi* vectors which have been reported in Kenya and the Horn of Africa [27]. In addition, artemisinin partial resistance has been reported in some parts of Kagera region near the border with Rwanda and these parasites can potentially spread to other regions if not curtailed (Giesbretch D and Ishengoma D, Unpublished data). Although recent studies have reported low prevalence of parasites with *hrp2/3 gene* deletions associated with failure of HRP2-based RDTs [14], these threats are still imminent and the country needs to increase its vigilance through a strong surveillance system. It is also critical to urgently adopt the WHO strategy for dealing with artemisinin partial resistance [28] to prevent spread of resistant parasites from Kagera to other regions.

This study assessed the prevalence and risk of malaria infections among asymptomatic individuals living in an area with high prevalence of parasites with artemisinin partial resistance in Kyerwa District, North-western Tanzania. Kyerwa is one of the districts of Kagera region with high malaria burden [29] and is located in an area which was recently reported to have high prevalence of artemisinin partial resistant parasites (Giesbretch D and Ishengoma D, Unpublished data). However, there is paucity of data regarding the magnitude and prevalence of infections in asymptomatic individuals, which serves a potential role in the transmission and risk of malaria infections, and potential spread of resistant parasites. Therefore, this study aimed at generating important data which will support the understanding of the burden and risk of malaria infections as well as the epidemiology of artemisinin partial resistant parasites. The findings will potentially help the government and other stakeholders in planning and implementing strategies for malaria prevention and control as well as mitigating artemisinin partial resistance in Kagera region.

## Methods

### Study design and sites

This was a community-based cross-sectional survey (CSS) which was conducted after the peak of transmission season from July to August 2023. It was part of the main project on molecular surveillance of malaria in Tanzania (MSMT) which has been running in 13 regions of Mainland Tanzania from 2021 and was recently extended to cover all 26 regions (Ishengoma – unpublished data). The study covered five villages (Kitoma, Kitwechenkura, Nyakabwera, Rubuga and Ruko) which are part of the longitudinal component of the MSMT that was initiated in 2022 to monitor the trends and patterns of malaria in areas with special features as described elsewhere (Ishengoma, unpublished data). The study villages are located in Kyerwa district which is one of the eight districts of Kagera region (Figure 1). Kyerwa district covers an area of 3,086 Km² and borders Uganda to the North, Rwanda to the West and Karagwe district to the South. The study villages are located near the Kagera river basin, which forms the boundary between Kyerwa and Rwanda (West) and Uganda (North). According to the 2022 national census, Kyerwa district had a population of 412,910 people, including 211,685 females and 201,225 males, with average household size of 4.3, and sex ratio of 95.

**Figure 1:**
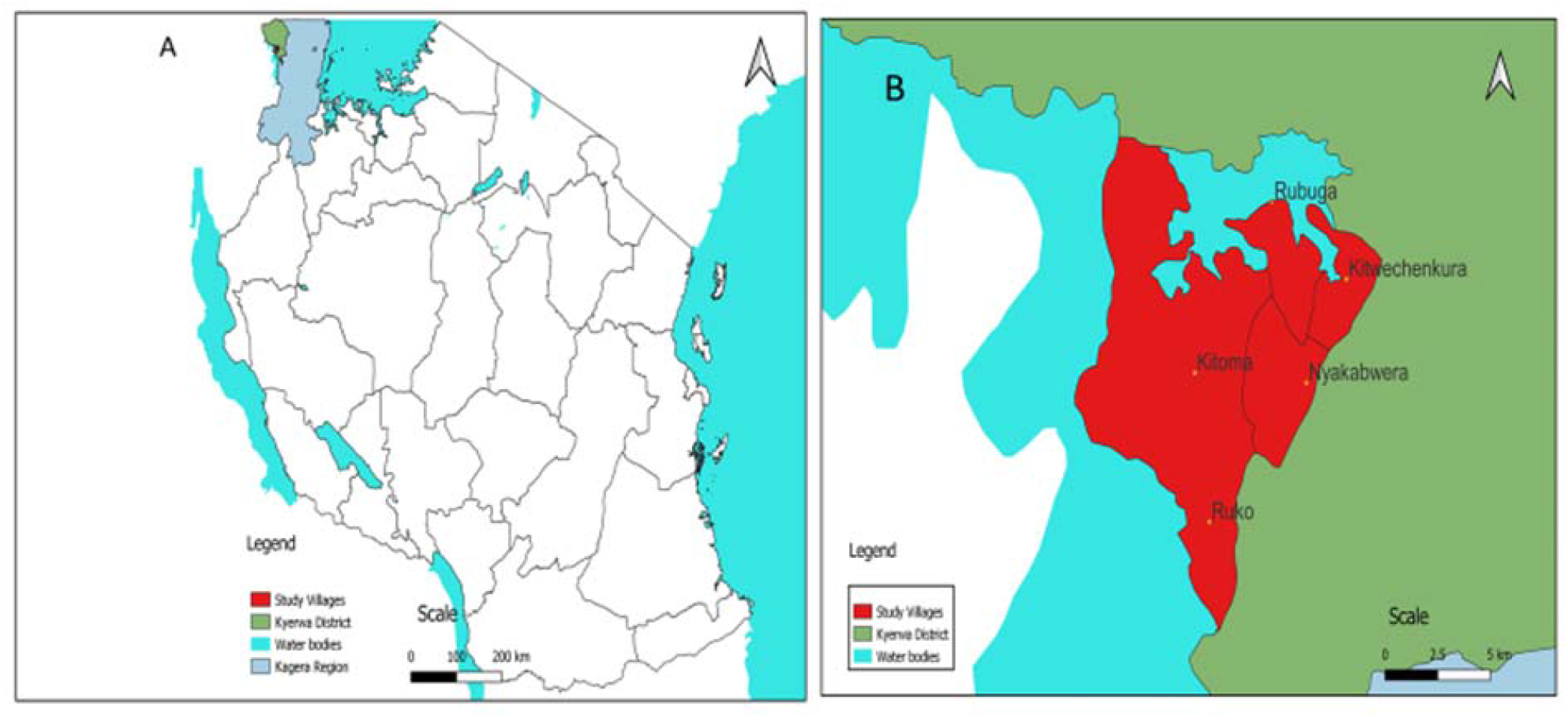
Maps show the study area; A: The 26 regions of Tanzania including Kagera (light blue) and Kyerwa district (green), B: Study villages in Kyerwa district (red).

### Study population and recruitment of participants

This CSS included individuals aged ≥6 months living in the five villages which are part of the longitudinal component of the MSMT project and provided consent to participate in the study. The survey targeted about 30% of all individuals from each village who were registered during a census survey that was conducted in February 2023. All community members meeting the inclusion criteria were invited to participant in the study. The inclusion criteria included age from 6 months and above, residence in one of the five villages and providing an informed consent. Individuals who wanted to participate but were residents of other villages or declined to give an informed consent were excluded from the CSS. A call for and information about the CSS was passed to all household members by a community organizer who broadcasted using a loudspeaker and walked through all parts of the village. Each of the study village had 3-5 hamlets and members from 1-3 hamlets were invited to meet the study team on specific days during the surveys. The study team was stationed at a central location that was set-up in each village. Any member who missed out was allowed to visit the survey location on a different day in the same or the next village.

### Data collection procedures

Data collected included demographic, anthropometric, clinical and parasitological data as shown in figure 2. Initially, basic demographic data were collected during the census survey that covered all households and individuals in each family. Every member of the community was enumerated during the census and given a unique identification number (IDs) and a database of all community members was set up at the National Institute for Medical Research (NIMR) in Dar es Salaam. During the CSS, each participant was identified using their IDs which are linked to the main database containing all members of the community. After confirming the identity of the prospective participants, they were provided with study identification numbers which were generated specifically for this CSS. Thereafter, participants were interviewed to obtain demographic information on top of those collected during the census. Each study participant was asked to report about malaria prevention practices through a series of questions, including how often they slept under a bed net (ITN or LLIN), how frequently other family members used bed nets, their habit of checking and repairing mosquito nets, and the regularity of using anti-mosquito spray/repellent or other methods within their household. They were later sent to designated areas for collection of anthropometric measurements (weight, height and body temperature) and then proceeded to the laboratory for testing for malaria and collection of blood samples. For detection of malaria parasites, participants were screened using RDTs (Abbott Bioline Malaria Ag Pf/Pan, Abbott Diagnostics Korea Inc., Korea, and Smart Malaria Pf/Pan Ag Rapid Test, Zhejiang Orient Gene Biotech Co. Ltd, China); and donated dried blood samples on filter paper (DBS) for laboratory analyses which will be reported elsewhere. Blood slides for detection of malaria parasites by microscopy were also collected for reading in the laboratory but the results are not included in this paper. The final step involved clinical assessment and collection of data on history of any illness as well as use of antimalarials and other medications (Fig. 2). Additional information including demographics, place of residence and presence of breeding sites were collected during the census survey from each household and all residents.

**Figure 2:**
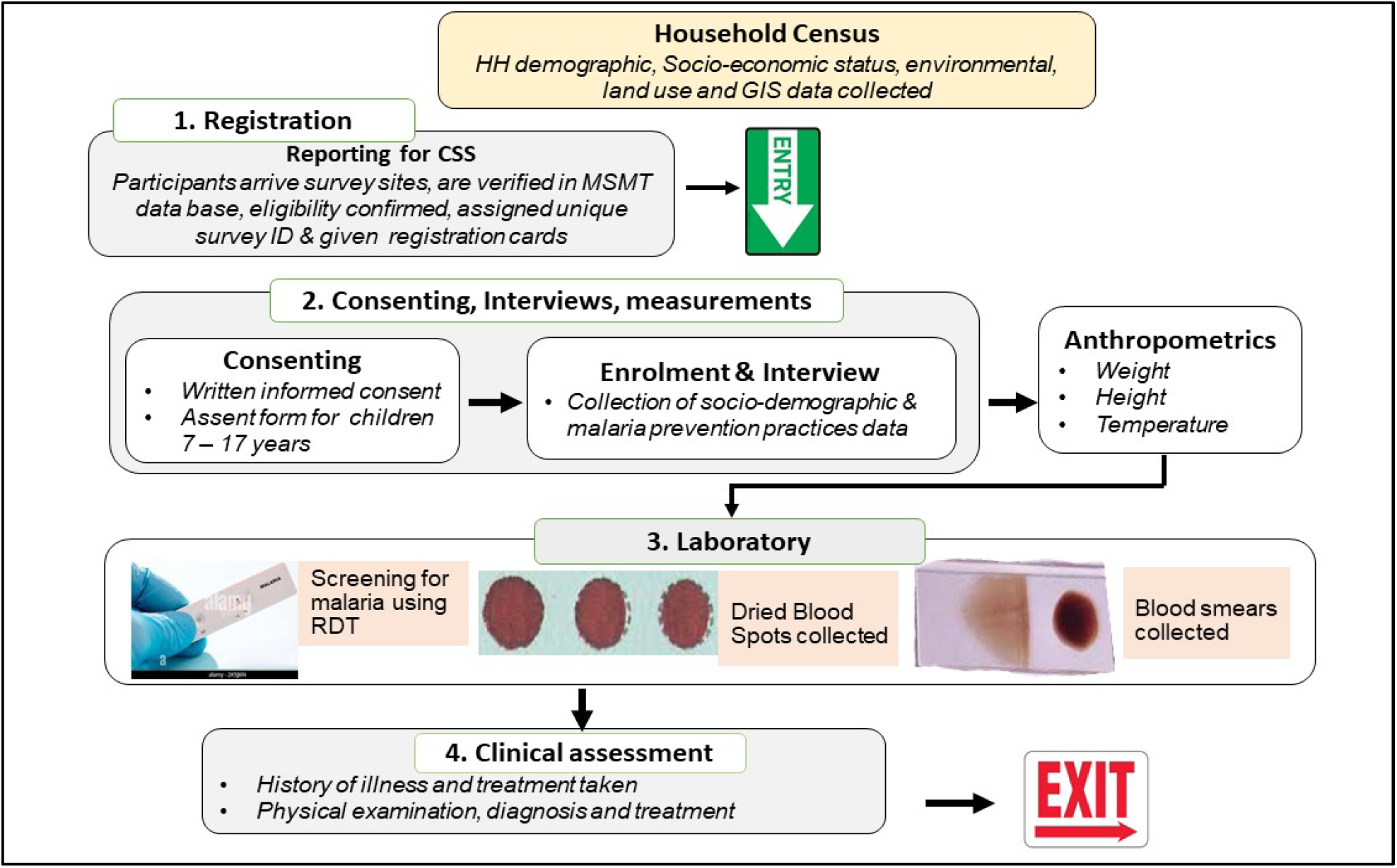
Schematic drawing showing the flow of study participants during the CSS Bed net use and other control interventions against malaria

This is the first CSS to be conducted in these villages based on the methods that have been used in previous studies conducted in selected communities of Tanga region from 1992 as described previously [30,31]. All asymptomatic (and some few symptomatic) participants were tested with RDTs and those with positive results were treated with Artemether-Lumefantrine and any other medicines according to the national guidelines [32]. Assessment of vector control methods was done by asking each participant to report about ownership of bed nets and their use as well as any information about the use of other methods. Through interviews with the district malaria focal person (DMIFP) and during the census survey, information about sources of ITNs/LLINs, use of other vector control interventions such IRS and LSM was collected. It was reported that Kyerwa receives bed-nets from NMCP through the different campaigns such as mass distribution, antenatal care (ANC) clinics and school net programme [33,34], together with private distributors. According to the NMCP reports of 2022, bed-net ownership in Kyerwa was at 89% and use was >80% (NMCP Unpublished data). The study villages and other parts of Kyerwa district and the entire region of Kagera were covered by the IRS programme funded by the US President’s Malaria Initiative (PMI) from 2008 to 2013 (Zilyahuramu G, Personal communication). The villages were also involved in LSM operations using biolarvicides produced in Tanzania but these activities could not be sustained because of lack of funding (Zilyahuramu G, Personal communication). Additional information about surveys of malaria vectors was explored from the DMIFP and other sources; indicating that the villages took part in entomological surveillance in the past.

### Data management and analysis

The data was collected with structured questionnaires created using ODK software, and installed and running on the tablets. The data was directly transmitted to a central server located at NIMR in Dar es Salaam. Data cleaning begun with exported the data to excel and daily cleaning which was done concurrently with field activities. Thereafter, the data were transferred to STATA version 13 (STATA Corp Inc., TX, USA) for further cleaning and analysis. Descriptive and statistical methods were used to describe the study population and generate baseline characteristics. The summary of results was presented in tables, figures, and text. Binary logistic regression was used to assess statistical association between malaria prevalence and different independent variables, before and after adjustments for covariates such as sex, age, age group and use of bed nets. The analysis also included other variables such as presence of breeding sites, types of houses and social economic status (SES) of the household. The household SES was calculated using assets owned by each household based on the data which were collected during the census survey in February 2023. Principal component analysis (PCA) was used to calculate SES quantiles using the data of household’s assets as previously described [35]. The items included in the PCA were ownership of a house, number of rooms, source of energy for lighting and cooking, source of drinking water, number of acres of land which was cultivated by the family, type of toilet, and possession of domestic animals and devices such as mobile phones, radio, television, bicycle, and motorcycle. Variables which were statistically significant (p-value <L0.25) during univariate logistic regression were included in multivariable logistic regression to control for the role of confounding variables. During the analysis, two separate models were used, the first included demographic and clinical variables, while the second model included household and environmental characteristics. P-value of ≤0.05 was considered to be significant.

## Results

### Baseline characteristics study participants

A total of 4,454 individuals constituting 29.9% of all community members provided consent and were enrolled in the CSS from the five villages (Kitoma, Kitwechenkura, Nyakabwera, Rubuga, and Ruko). Most of the participants were females (59.3%) and their median age was 14 years (interquartile range - IQR: 7 – 36 years), with significant differences in the age of participants among the study villages (p<0.01). Majority of the participants were students/children (49.5%) and (48.8 %) were peasants. Out of the adults, about 42.8% had completed primary education while (30.5%) did not have formal education. About 30% of the participants had a history of fever in the past 48 hours before the survey, but only 3.1% had fever at presentation (with axillary temperature ≥37.5^0^C) (Table 1).

**Table 1:**
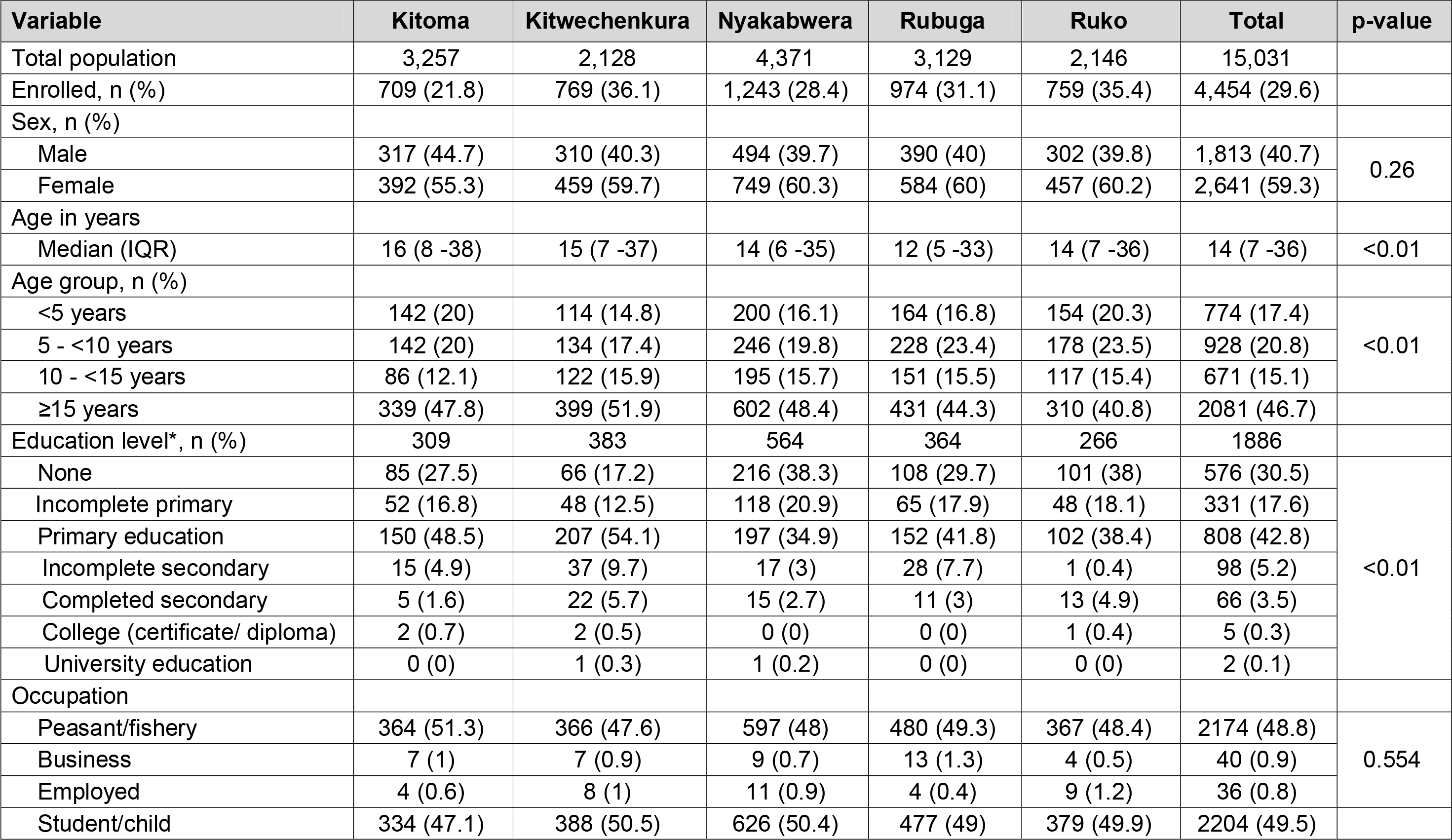
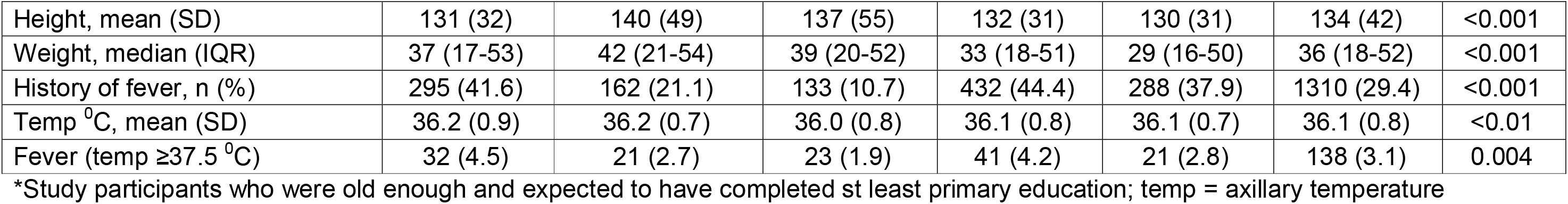
Baseline characteristics of study participants from the five village of Kyerwa district of Kagera region.

| Variable | Kitoma | Kitwechenkura | Nyakabwera | Rubuga | Ruko | Total | p-value |
| --- | --- | --- | --- | --- | --- | --- | --- |
| Total population | 3,257 | 2,128 | 4,371 | 3,129 | 2,146 | 15,031 |  |
| Enrolled, n (%) | 709 (21.8) | 769 (36.1) | 1,243 (28.4) | 974 (31.1) | 759 (35.4) | 4,454 (29.6) |  |
| Sex, n (%) |  |  |  |  |  |  |  |
| Male | 317 (44.7) | 310 (40.3) | 494 (39.7) | 390 (40) | 302 (39.8) | 1,813 (40.7) | 0.26 |
| Female | 392 (55.3) | 459 (59.7) | 749 (60.3) | 584 (60) | 457 (60.2) | 2,641 (59.3) |  |
| Age in years |  |  |  |  |  |  |  |
| Median (IQR) | 16 (8 -38) | 15 (7 -37) | 14 (6 -35) | 12 (5 -33) | 14 (7 -36) | 14 (7 -36) | <0.01 |
| Age group, n (%) |  |  |  |  |  |  |  |
| <5 years | 142 (20) | 114 (14.8) | 200 (16.1) | 164 (16.8) | 154 (20.3) | 774 (17.4) | <0.01 |
| 5 - <10 years | 142 (20) | 134 (17.4) | 246 (19.8) | 228 (23.4) | 178 (23.5) | 928 (20.8) |  |
| 10 - <15 years | 86 (12.1) | 122 (15.9) | 195 (15.7) | 151 (15.5) | 117 (15.4) | 671 (15.1) |  |
| ≥15 years | 339 (47.8) | 399 (51.9) | 602 (48.4) | 431 (44.3) | 310 (40.8) | 2081 (46.7) |  |
| Education level*, n (%) | 309 | 383 | 564 | 364 | 266 | 1886 |  |
| None | 85 (27.5) | 66 (17.2) | 216 (38.3) | 108 (29.7) | 101 (38) | 576 (30.5) | <0.01 |
| Incomplete primary | 52 (16.8) | 48 (12.5) | 118 (20.9) | 65 (17.9) | 48 (18.1) | 331 (17.6) |  |
| Primary education | 150 (48.5) | 207 (54.1) | 197 (34.9) | 152 (41.8) | 102 (38.4) | 808 (42.8) |  |
| Incomplete secondary | 15 (4.9) | 37 (9.7) | 17 (3) | 28 (7.7) | 1 (0.4) | 98 (5.2) |  |
| Completed secondary | 5 (1.6) | 22 (5.7) | 15 (2.7) | 11 (3) | 13 (4.9) | 66 (3.5) |  |
| College (certificate/ diploma) | 2 (0.7) | 2 (0.5) | 0 (0) | 0 (0) | 1 (0.4) | 5 (0.3) |  |
| University education | 0 (0) | 1 (0.3) | 1 (0.2) | 0 (0) | 0 (0) | 2 (0.1) |  |
| Occupation |  |  |  |  |  |  |  |
| Peasant/fishery | 364 (51.3) | 366 (47.6) | 597 (48) | 480 (49.3) | 367 (48.4) | 2174 (48.8) | 0.554 |
| Business | 7 (1) | 7 (0.9) | 9 (0.7) | 13 (1.3) | 4 (0.5) | 40 (0.9) |  |
| Employed | 4 (0.6) | 8 (1) | 11 (0.9) | 4 (0.4) | 9 (1.2) | 36 (0.8) |  |
| Student/child | 334 (47.1) | 388 (50.5) | 626 (50.4) | 477 (49) | 379 (49.9) | 2204 (49.5) |  |
| Height, mean (SD) | 131 (32) | 140 (49) | 137 (55) | 132 (31) | 130 (31) | 134 (42) | <0.001 |
| Weight, median (IQR) | 37 (17-53) | 42 (21-54) | 39 (20-52) | 33 (18-51) | 29 (16-50) | 36 (18-52) | <0.001 |
| History of fever, n (%) | 295 (41.6) | 162 (21.1) | 133 (10.7) | 432 (44.4) | 288 (37.9) | 1310 (29.4) | <0.001 |
| Temp °C, mean (SD) | 36.2 (0.9) | 36.2 (0.7) | 36.0 (0.8) | 36.1 (0.8) | 36.1 (0.7) | 36.1 (0.8) | <0.01 |
| Fever (temp ≥37.5 °C) | 32 (4.5) | 21 (2.7) | 23 (1.9) | 41 (4.2) | 21 (2.8) | 138 (3.1) | 0.004 |

\*Study participants who were old enough and expected to have completed st least primary education; temp = axillary temperature

### Prevalence of malaria

All individuals (n=4,454) were tested with RDTs and 1,979 (44.4%) had positive results. The prevalence of malaria in the five villages varied from 14.5% in Nyakabwera to 68.5% in Ruko village suggesting a highly heterogeneous transmission in these villages which are located next to each other. Higher prevalence was detected in males (49.2%) in all villages except in Kitoma, and in school children (aged 5 - ≤ 10 years with 61.1% and 10 - <15 years with 57.7%). The lowest prevalence was in adults aged ≥15 years (29.5%), and the pattern of age-specific prevalence was similar in all villages (Table 2). In all villages except Nyakabwera (p=0.07), the prevalence of malaria was significantly higher in school children (p<0.01). Majority of the individuals with malaria infections were clustered near water bodies and other types of breeding sites in all villages (Figure 3).

**Figure 3:**
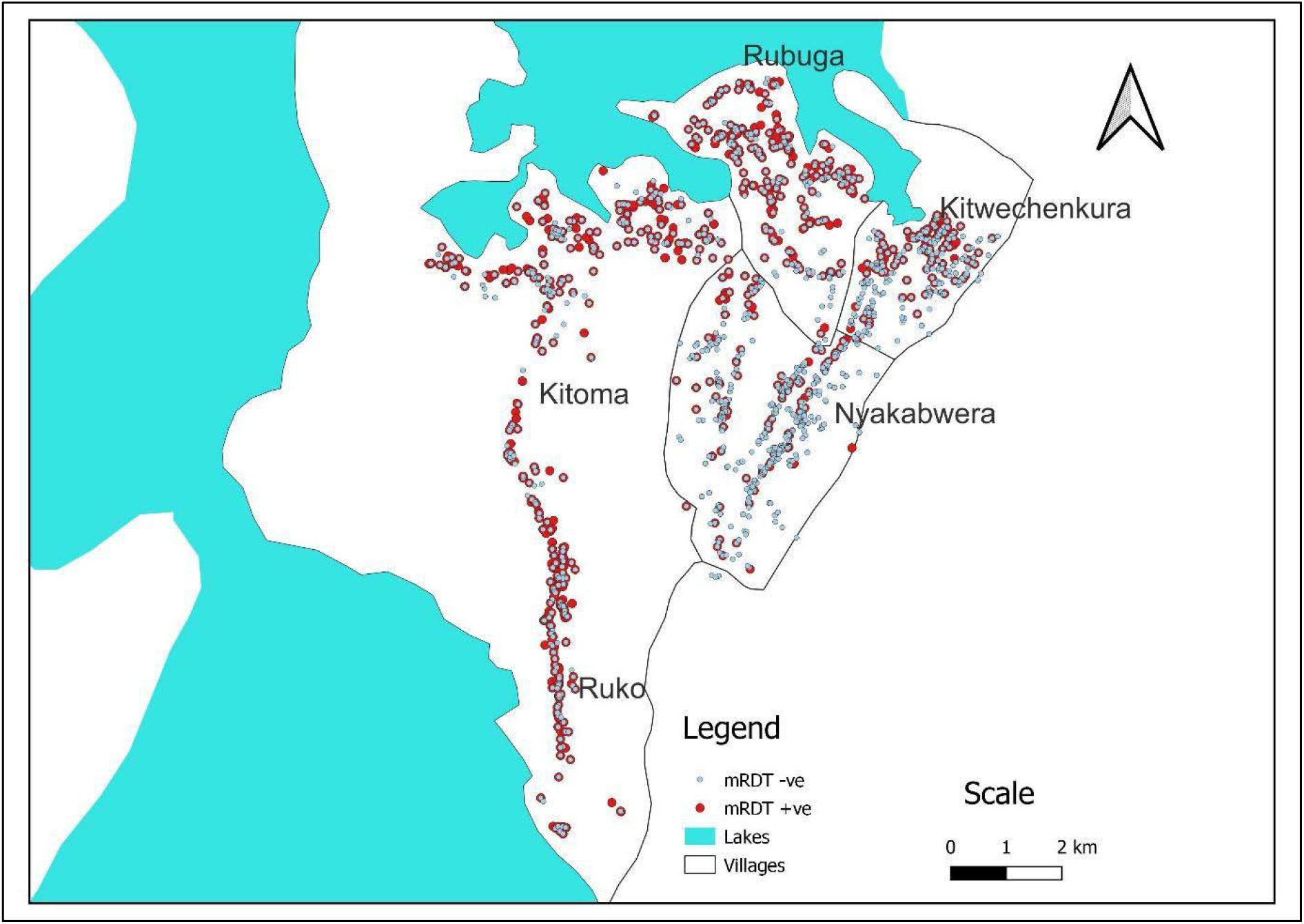
Map of study villages showing clustering of individuals with a positive test for malaria

**Figure 4:**
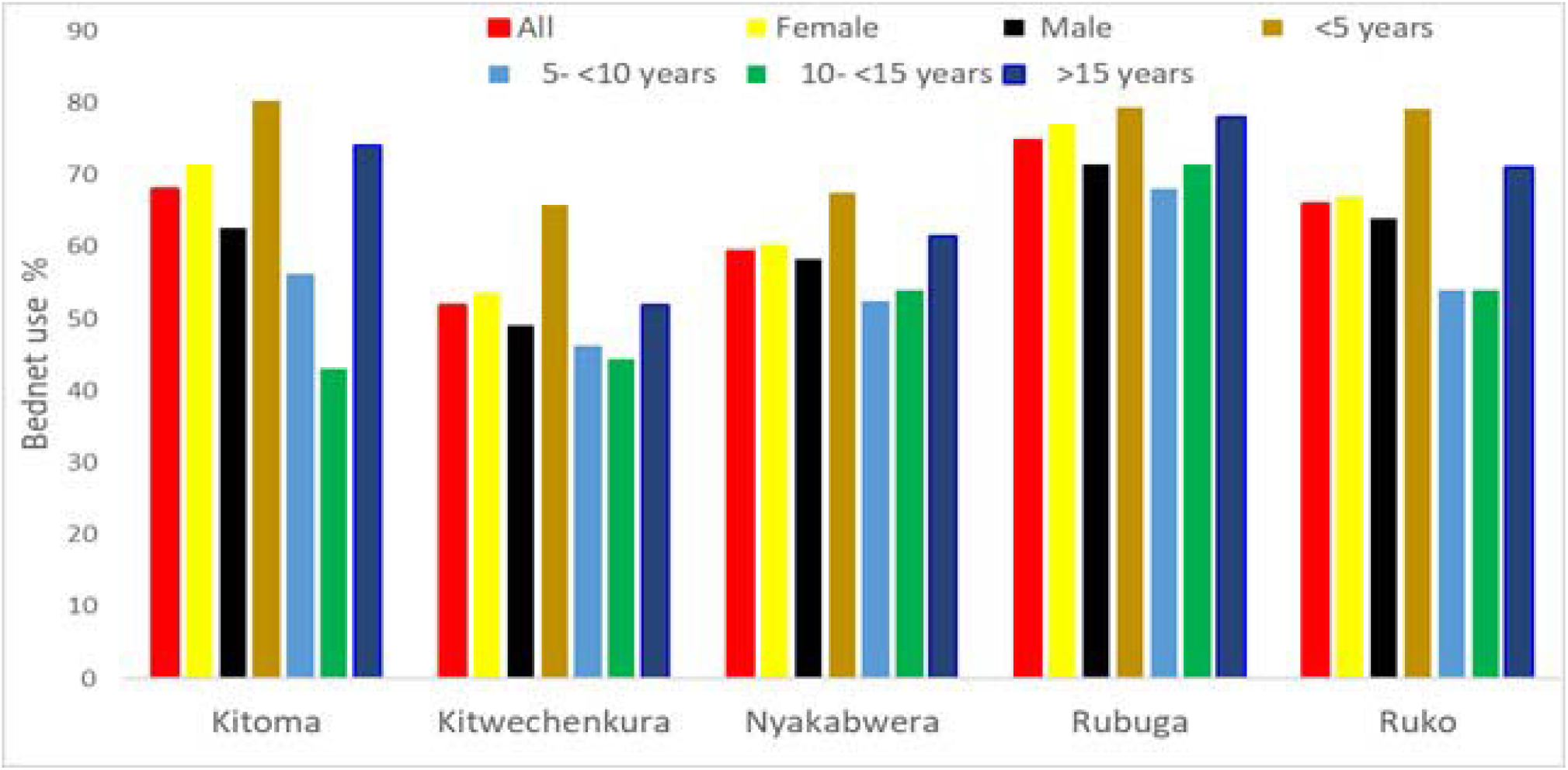
Bed-net usage by sex and age in the five villages of Kyerwa

**Table 2:**
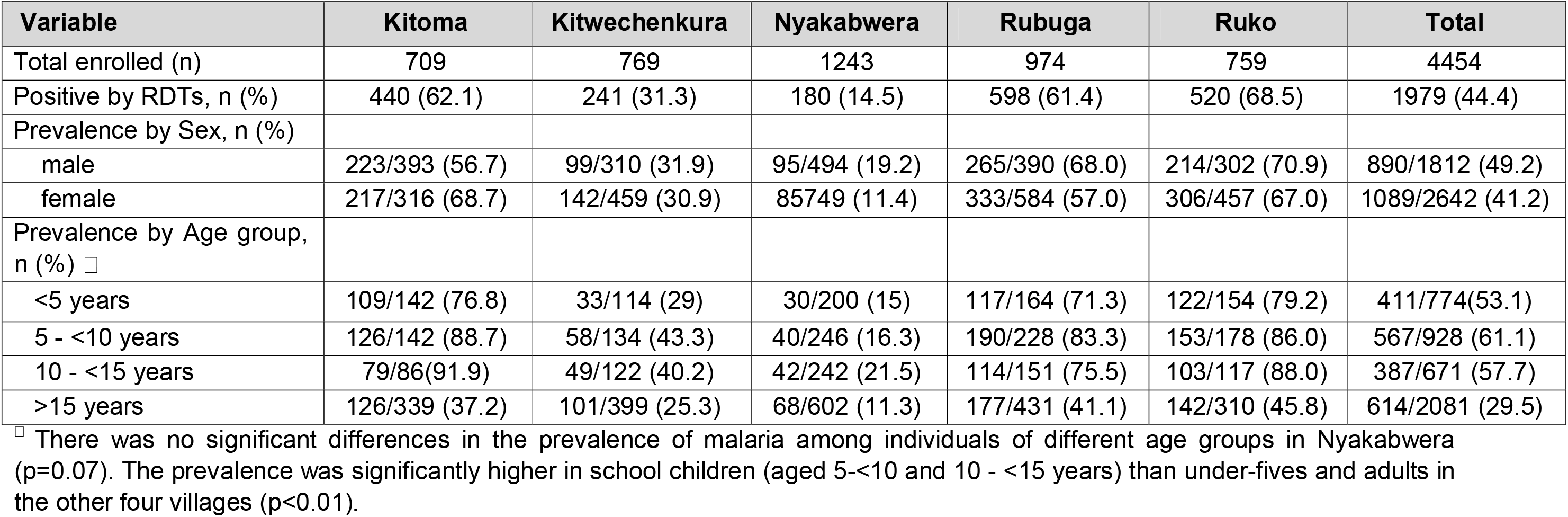
Prevalence of malaria in the five villages of Kyerwa.

### Bed net use in the study villages

Table 3 shows the use of bed nets and other methods of preventing malaria among study participants. The overall use of bed nets in the past night before the survey (in all villages combined) was 63.9%, with significantly higher usage among females (65.7%) compared to males (61.5%) in all villages combined (p = <0.01); and in each of the villages (p≤0.02). Bed net usage was higher in Rubuga (74.5%) and Kitoma (68.0%) while the lowest was reported in Kitwechenkura (51.8%). There was low bed net usage among school children (56.3% and 54.7% in children aged 5 - <10 and 10 - <15 years, respectively) compared to under-fives and adults (≥15 years) with usage of ≥67.0% (Table 3). In all five villages, females, under-fives and adults (≥ 15 years) had higher usage of nets, with three villages (Kitoma, Ruko and Rubuga) approaching or exceeding 80% (Figure 3). Only a small proportion of the participants (2.4%) reported use of other methods to protect themselves against mosquito bites (Table 3). Such methods included mosquito repellents (n=67, 1.5%), mosquito coils (n=21, 0.5%) and burning insecticides (n=18, 0.4%).

**Table 3:**
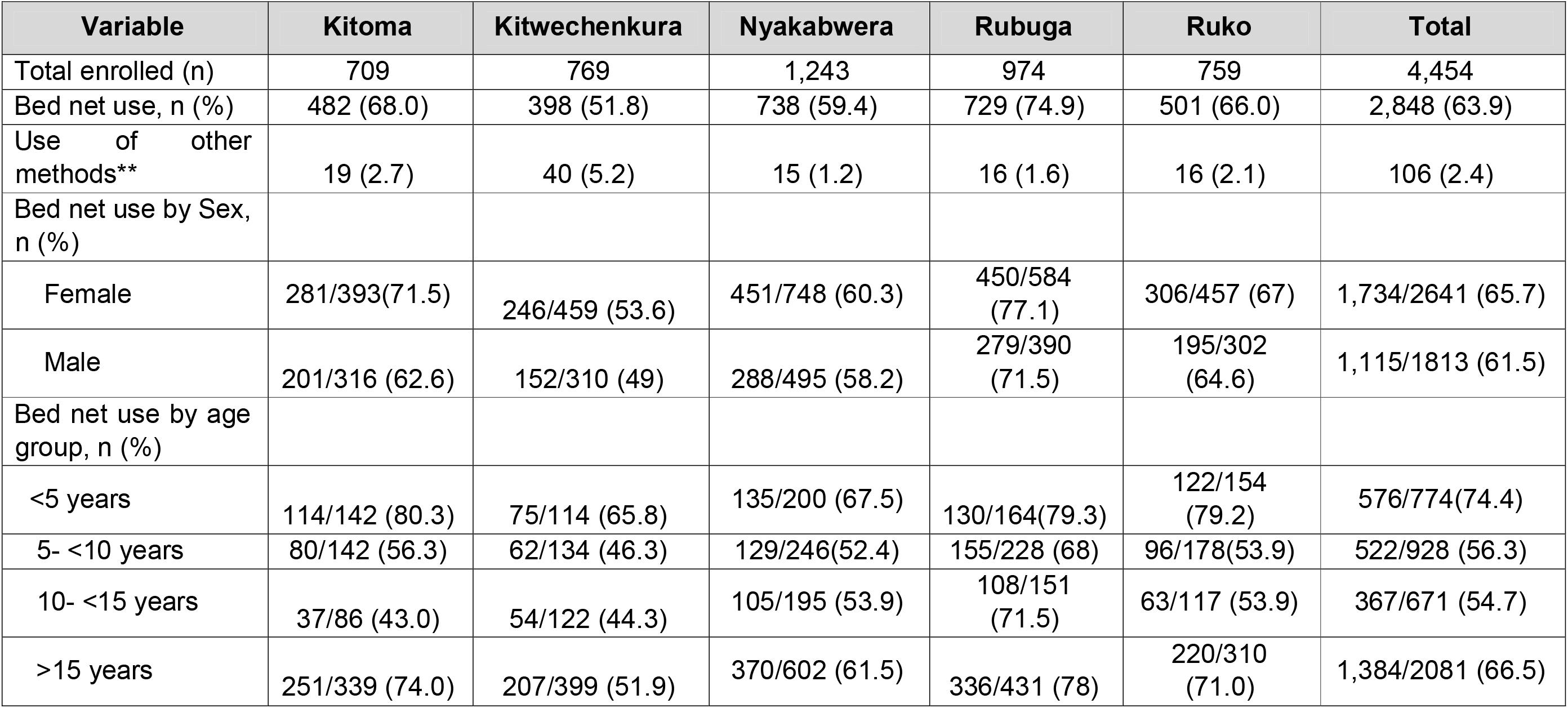
Use of bed-nets among study participants.

### Risk factors of malaria infections

When compared to Nyakabwera, the risk of malaria infections was significantly higher in the other four villages; with three times higher risk in Kitwechenkura and over 11.0 times in the three villages of Kitoma, Ruko and Rubuga (p<0.001). The risk was 19.0% higher in males compared to females (p= 0.017) while individuals who reported that they did not use bed nets had 27.0% more risk compared to those who used nets a night before the survey (p= 0.002). School children and under-fives had significantly higher risk of malaria infections (p<0.001) compared to adults (≥15 years). The risk of malaria infection was 22 times among participants with a history of fever in the past two days before the survey (p<0.001) (Table 4a). Individuals with low SES had higher risk of malaria infection by 27% (p = 0.020) compared to those with high SES. Living in houses with open windows (p=0.024) and walls made of mud (p<0.001) were associated with higher risk of malaria infections (Table 4b).

**Table 4a:**
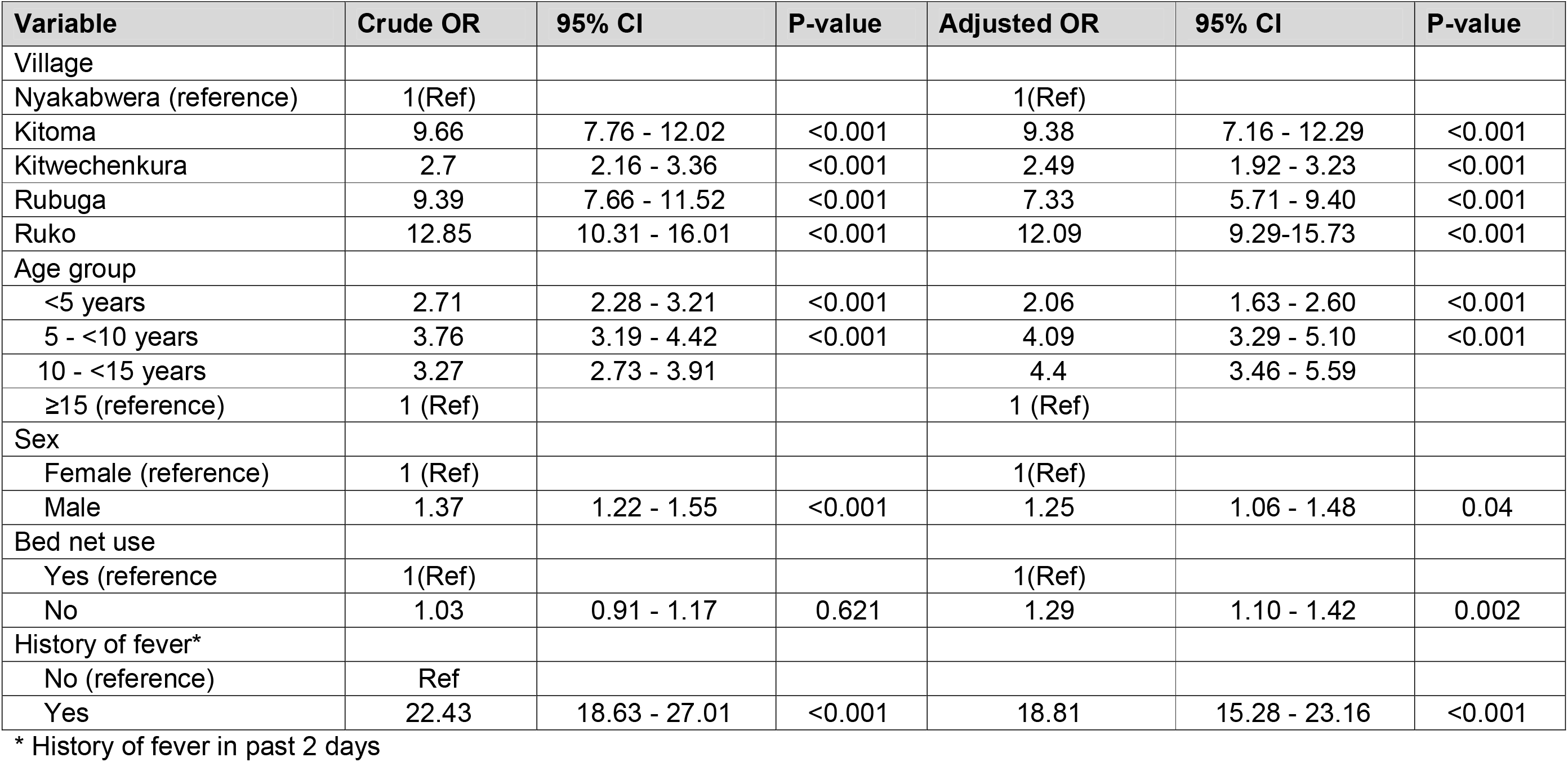
Demographic and clinical factors associated with malaria infections in the five villages of Kyerwa.

| Variable | Crude OR | 95% CI | P-value | Adjusted OR | 95% CI | P-value |
| --- | --- | --- | --- | --- | --- | --- |
| Village |  |  |  |  |  |  |
| Nyakabwera (reference) | 1(Ref) |  |  | 1(Ref) |  |  |
| Kitoma | 9.66 | 7.76 - 12.02 | <0.001 | 9.38 | 7.16 - 12.29 | <0.001 |
| Kitwechenkura | 2.7 | 2.16 - 3.36 | <0.001 | 2.49 | 1.92 - 3.23 | <0.001 |
| Rubuga | 9.39 | 7.66 - 11.52 | <0.001 | 7.33 | 5.71 - 9.40 | <0.001 |
| Ruko | 12.85 | 10.31 - 16.01 | <0.001 | 12.09 | 9.29-15.73 | <0.001 |
| Age group |  |  |  |  |  |  |
| <5 years | 2.71 | 2.28 - 3.21 | <0.001 | 2.06 | 1.63 - 2.60 | <0.001 |
| 5 - <10 years | 3.76 | 3.19 - 4.42 | <0.001 | 4.09 | 3.29 - 5.10 | <0.001 |
| 10 - <15 years | 3.27 | 2.73 - 3.91 |  | 4.4 | 3.46 - 5.59 |  |
| ≥15 (reference) | 1 (Ref) |  |  | 1 (Ref) |  |  |
| Sex |  |  |  |  |  |  |
| Female (reference) | 1 (Ref) |  |  | 1(Ref) |  |  |
| Male | 1.37 | 1.22 - 1.55 | <0.001 | 1.25 | 1.06 - 1.48 | 0.04 |
| Bed net use |  |  |  |  |  |  |
| Yes (reference) | 1(Ref) |  |  | 1(Ref) |  |  |
| No | 1.03 | 0.91 - 1.17 | 0.621 | 1.29 | 1.10 - 1.42 | 0.002 |
| History of fever* |  |  |  |  |  |  |
| No (reference) | Ref |  |  |  |  |  |
| Yes | 22.43 | 18.63 - 27.01 | <0.001 | 18.81 | 15.28 - 23.16 | <0.001 |

\* History of fever in past 2 days

**Table 4b:** Household and environmental characteristics associated with malaria infections in the five villages in Kyerwa districts.

| Variable | Crude OR | 95% CI | P-value | Adjusted OR | 95% CI | P-value |
| --- | --- | --- | --- | --- | --- | --- |
| Family size |  |  |  |  |  |  |
| <4 people (reference) | Ref |  |  |  |  |  |
| >4 people | 1.11 | 0.98 - 1.26 | 0.104 |  |  |  |
| Wall type (reference) |  |  |  |  |  |  |
| Blocks/bricks | Ref |  |  |  |  |  |
| Mud | 0.67 | 0.59 - 0.75 | <0.001 | 1.61 | 1.23 - 2.09 | <0.001 |
| Roof type |  |  |  |  |  |  |
| Metal (reference) | Ref |  |  |  |  |  |
| Thatch | 1.27 | 0.95 - 1.69 | 0.104 |  |  |  |
| Floor type |  |  |  |  |  |  |
| Cement (reference) | Ref |  |  |  |  |  |
| Sand/soil | 1.41 | 1.23 - 1.60 | <0.001 | 1.19 | 0.99-1.43 | 0.058 |
| SES |  |  |  |  |  |  |
| High (reference) | Ref |  |  |  |  |  |
| Low | 1.64 | 1.41 - 1.92 | <0.001 | 1.27 | 1.04-1.55 | 0.02 |
| Medium | 1.07 | 0.93 - 1.23 | 0.35 | 0.97 | 0.84 - 1.20 | 0.972 |
| Type of eaves |  |  |  |  |  |  |
| Closed (Reference) | Ref |  |  | Ref |  |  |
| Open | 1.21 | 1.07 - 1.38 | 0.004 | 1.06 | 0.89 -1.25 | 0.533 |
| Type of windows |  |  |  |  |  |  |
| Closed (reference) | Ref |  |  | Ref |  |  |
| Open | 1.2 | 1.02 - 1.42 | 0.024 | 1.26 | 1.03 - 1.54 | 0.024 |
| Partially open | 1.2 | 0.93 - 1.56 | 0.168 | 1.32 | 0.96 - 1.82 | 0.085 |
| No window | 1.97 | 1.42 - 2.74 | <0.001 | 1.38 | 0.92 - 2.08 | 0.12 |
| Vegetation cover near the houses |  |  |  |  |  |  |
| No (reference) | Ref |  |  | Ref |  |  |
| Short vegetation | 1.33 | 1.05 - 1.68 | 0.017 | 1.11 | 0.83 -1.47 | 0.492 |
| Long vegetation | 1.63 | 1.30 - 2.05 | <0.001 | 1.28 | 0.96 -1.70 | 0.088 |
| Presence of bushes around the houses |  |  |  |  |  |  |
| No (reference) | Ref |  |  |  |  |  |
| Yes | 1.29 | 1.14 - 1.46 | <0.001 | 1.04 | 0.87 - 1.23 | 0.682 |

## Discussion

This CSS was conducted as part of a larger project on MSMT which aims at establishing the capacity and implementing malaria molecular surveillance (MMS) in Tanzania [36]. The MSMT project has been implemented in 13 regions of Tanzania from 2021 and it has now been extended to cover integrated malaria molecular surveillance (iMMS) in all 26 regions of Mainland Tanzania, from January 2023 (Ishengoma et al. Unpublished data). This study generated important baseline data in five villages which are located in an area with high prevalence of artemisinin partial resistance in Kagera region, North-western Tanzania (Giesbrecht D and Ishengoma D, Unpublished data). The study utilised a platform which has been set-up by the MSMT project to support studies focused on iMMS in Tanzania. It enrolled asymptomatic individuals from the five villages under the longitudinal surveillance component of MSMT and showed that all villages had a very high prevalence of malaria infections by RDTs (44.4%), with high variability despite their close proximity. In these villages, the risk of malaria infections was significantly higher in males, school children and individuals with low SES, a history of fever (within two days before the CSS) as well as those who were not using bed nets. In all villages, the prevalence and risk of malaria infections were heterogeneous at micro-geographic levels, despite high use of bed-nets (over 51%) which exceeded 66% in three villages. The findings reported here will be critical in future studies to characterise and monitor transmissibility, trends and patterns as well as spread of artemisinin partial resistant parasites in the region and other parts of Tanzania.

In this study, the majority of participants were females compared to males and most of them were peasants or fishermen. In addition, the majority (about 70%) were either illiterate, did not complete or had primary education, and over 26% were school children. These demographic features present a population of individuals with potentially high risk of malaria due to low SES (shown to be associated with high risk of malaria infections in this study). Preliminary analysis of the census data collected in this community showed that the majority of the people with low SES had similar features; which included being females, reporting low education level, and were peasants (Challe, unpublished data). Studies conducted elsewhere also reported that women were more likely to take part in research studies due to their role as care-givers and perceived high risk of malaria compared to males [37]. In previous studies, it was also shown that low SES was an important risk factor for malaria infections [35,38–40], mainly due to the strong association between malaria and poverty [41,42]. Poor people have higher risk of malaria infections because they cannot afford the cost of malaria prevention and/or treatment and also lack or have low level of knowledge and skills required to protect themselves against malaria infections [43].

Although this study targeted asymptomatic individuals from the community, about 29.0% of the participants reported that they had a history of fever in the past 48 hours and only 3.1% had fever at presentation (axillary temperature ≥37.5^0^C). This could be possibly due to over-reporting by study participants in anticipation of getting medicines for malaria (C. Mandara et al. Personal Communication). In our previous studies, it was observed that study participants reported to have malaria or any other febrile illnesses because they wanted to get better services from the study team and medicines which were given for free. It was also shown that participants would prefer to be given medicines to keep for future use or to share with family members who were not present at the time of the study or in case they fell sick. When data analysis was done to tease out the relationship between reported fever history and the results of RDTs, there was a significant association between history of fever and malaria infection [44,45] but this could be due high prevalence of malaria in study villages. Thus, fever history was possibly over-reported by participants in anticipation of getting better services and medicines. Similarly, there was a strong association between fever at presentation (axillary temperature ≥37.5^0^C) and malaria infection and this was similar to the findings of other community studies which showed that fever at presentation was a strong predictor of malaria infection[46].

The overall malaria prevalence in this community was very high (over 44%), with higher prevalence among males, and in three of the five villages, the prevalence exceeded 61.0%. There was very high heterogeneity across the five villages which are located close to each other, suggesting that there are critical factors associated with the micro-geographic pattern of malaria in this community. The findings are similar to what was reported elsewhere [47], where high level of heterogeneity at micro-geographic levels was reported within the wards from 80 councils of Mainland Tanzania. Although few studies have assessed the prevalence of malaria in asymptomatic individuals of all age groups in Tanzania, the prevalence reported in this study is likely the highest in the country in recent years. Studies conducted in Tanga reported that the prevalence of malaria among under-fives and school children were higher (≥68.0%) between the year 1992 and 1999 but declined to less than 10% in 2012 [48]. Following an increase of malaria in Tanga, the prevalence increased but the highest was 31.4% in 2015 [31]. In Rufiji, a decline of prevalence was also reported and the highest was 90.0% in 1985 while the surveys conducted between 2001 and 2006 reported the highest prevalence of 23% [49]. The causes of this high level of prevalence in these villages of Kyerwa district need to be established to guide specific interventions to reduce the burden of malaria which will potentially reduce the spread of drug resistant parasites currently circulating in this and other areas of Kagera region.

The age specific prevalence showed significantly higher prevalence in school children (aged 5 - <15 years) followed by under-fives while adults (≥15 years) had the lowest prevalence (overall and in each of the five villages). High prevalence of malaria among school children has consistently been reported in studies which were conducted in Tanga from 2008 [31]. A similar trend has been reported in other parts of Tanzania particularly through the school surveys. Over the past 10 years, high prevalence of malaria among school children has been reported with prevalence reaching 76.4% in some district councils [21]. However, a declining trend of malaria in school children has been observed in the school surveys which were conducted from 2015 to 2021, with an overall decline from 21.7% in 2015 to 11.3% in 2021 (Chacky *et al*. unpublished data). In this study, the observed high prevalence of malaria among school children which was ≥75.5% and reached 92% in the three villages with high prevalence (Kitoma, Rubuga and Ruko) needs to be monitored to determine the factors associated with this high burden of malaria in this group. Future studies will also need to focus on the potential lack or low impact of different interventions against malaria in this community and specifically those directed at school children.

Three of the five villages had high bed net use (over 66.0%) while two villages had low use (less than 60%) and one village had very low use of bed nets (51.9%). In all villages, bed net use was higher among females compared to males, and was significantly lower in school children (overall and in each of the five villages). High bed net use among females is possibly due to the ongoing campaigns to provide free bed nets to pregnant women at ANC visits in Tanzania [34]. However, low bed net use by school children could account for the high prevalence of malaria in this group and this is surprising because a school net programme has been running in Tanzania since 2013 [50]. This campaign is aimed at controlling and reducing the burden of malaria in these children. Although previous studies reported an increase in bed net ownership among school children [33,51,52], it is possible that ownership could be high but the bed nets are not routinely used. Additional studies are needed to tease out the level of bed net ownership and use in this community and their impacts against malaria burden since school children who were expected to be protected by nets still had low use and the highest prevalence of malaria.

In all villages, under-fives had significantly higher bed-net use, followed by adults. In three of the five villages, use of bed nets among under-fives was closer to or exceeded 80% which is the cut-off recommended by WHO. Yet, under-fives had higher prevalence of malaria in all villages, although it was lower than that observed in school children. It was also shown that a high proportion of adults were using bed-nets and together with high bed net use among under-fives, this could be possibly due to free bed nets given to children and pregnant women attending ANC clinics in Tanzania [53]. However, the lack of impact of high bed net use in young children in this community needs to be further explored. Previous studies emphasised the importance of behaviour change in assessing malaria prevalence, specifically in relation to bed net coverage and use, because ownership may possibly not be directly related to use of nets. In addition, it has been shown that outdoor biting mosquitoes are the main cause of infections even in areas with high coverage and use of nets, especially when humans spend time outdoors during the early hours of the night [54]. Therefore, it is crucial to investigate the prevalence of outdoor biting mosquitoes and human behaviour that may expose individuals to malaria infection when outdoor or indoor but not sleeping under bed nets.

The findings of this study showed that the risk of malaria infections was higher in all villages compared to Nyakabwera, among males compared to females and in under-fives and school children compared to adults. It was also shown that individuals who used bed nets had lower risk (those who were not using nets had high risk by over 27% compared to bed net users). Additional risk factors for malaria infections included low SES, living in houses whose walls are made of muds and houses with open windows. As shown in previous studies, low SES and poor houses or living conditions are strongly associated with higher risk of malaria infections [35,38–40]. This suggests that efforts to eliminate malaria need to target groups at high risk, and to consider and concurrently implement strategies for poverty reduction. Although previous studies showed high heterogeneity of malaria transmission and risk at macro-geographic levels [47,55], there is paucity of studies of micro-geographic variations of malaria burden and associated risk factors [47]. Thus, more studies are required to further assess malaria burden and risk factors associated with its transmission at different geographic levels. Additional studies will also be required to determine if such high transmission will support the spread of resistant parasites in this and other areas of Kagera region and beyond.

## Conclusion

This study showed high prevalence of malaria infections and high heterogeneity at micro-geographic levels. The risk of malaria infections was higher in school children, males and individuals who reported that they did not use bed nets a night prior to the study as well as those with low SES, and living in poorly constructed houses. These findings provide important baseline data in an area with high prevalence of artemisinin partial resistant parasites and will be utilised in future studies to monitor the trends and potential spread of such parasites.

## Data Availability

The data used in this paper are available and can be obtained upon a request from the corresponding author.

## List of abbreviations

ACT: Artemisinin-based combination therapy
ANC: Antenatal care clinic
aOR: adjusted odds ratio
CI: Confidence interval
CSS: Cross-sectional community survey
DMFP: District malaria focal person
IDs: Identification numbers
iMMS: Integrated malaria molecular surveillance
IPTp: Intermittent preventive treatment
IRS: Indoor residual spraying
ITN: Insecticides treated Nets
IQR: Interquartile range
LSM: Larval source management
LLINs: Long-lasting insecticidal nets
MMS: Malaria molecular surveillance
MRCC: Medical Research Coordinating Committee
MSMT: Molecular surveillance of malaria in Tanzania.
RDTs: Rapid diagnostic tests for malaria
NIMR: National Institute for Medical Research
NMCP: National Malaria Control Program
NMSP: National malaria strategic plan
OR: Odds ratio
ODK: Open Data Kit
PO-RALG: President’s Office, Regional Administration and Local Government
PCA: Principal component analysis
SES: Socio-economic status
SP: Sulphadoxine-pyrimethamine TMIS Tanzania malaria indicator survey WHO World Health Organization
WHO-Afro: WHO Regional office for African

## Declarations

### Ethics approval and consent to participate

This CSS was part of the MSMT project whose protocol was reviewed and approved by the Medical Research Coordinating Committee (MRCC) of the National Institute for Medical Research (NIMR). Permission to conduct the study was sought from the President’s Office, Regional Administration and Local Government (PO-RALG), regional authorities of Kagera and the District Executive Director of Kyerwa. Leaders of Kitwechenkura ward and the five villages were consulted through meetings which were held at the ward level and agreed for this and other components of the MSMT project to be implemented in their communities. Information about the CSS was given in the community through their village mobilization teams for two consecutive days before the survey. Before taking part in the survey, verbal and written informed consent were sought and obtained from all participants or parents/guardians in case of children.

### Authors contribution

DSI conceived of the idea, supervised implementation of the study and data analysis, and was involved in the interpretation of the results. FF, DPC and DAP were involved in data collection and analysis and interpretation of the results. DSI and SSM, wrote the manuscript and all authors contributed to the final version. DSI revised and finalised the manuscript and all authors read and approved the manuscript.

## Acknowledgements

The study team is grateful to participants for their willingness to participate in the CSS and providing consent as well as taking part in the study. The contribution of the data collection and laboratory teams is highly appreciated. Those who participated in data collection and/or laboratory processing of samples included, Ezekiel Malecela, Oswald Oscar, Ildephonce Mathias, Gerion Gaudin, Kusa Mchaina, Hussein Semboja, Sharifa Hassan, Salome Simba, Hatibu Athumani, Ambele Lyatinga, Honest Munishi, Anael Derrick Kimaro, Ally Idrissa and Amina Ibrahim. Special thanks to the finance, administrative, and logistic support teams at NIMR: Christopher Masaka, Millen Meena, Beatrice Mwampeta, Neema Manumbu, Arison Ekoni, Sadiki Yusuph, John Fundi, Fred Mashanda, Amir Tununu and Andrew Kimboi. The support from the Management of NIMR, NMCP and PO-RALG was critical for the success of this CSS and it is therefore appreciated. The team appreciates technical and logistics support from partners at Brown University, the University of North Carolina at Chapel Hill, CDC Foundation and from the Bill and Melinda Gates Foundation team.

